# The perception and preference for online education among final year undergraduates in selected state universities in Western Province, Sri Lanka, during the COVID-19 Pandemic

**DOI:** 10.1101/2025.10.18.25338275

**Authors:** S. H. D. Upeksha, M. A. M. N. Gamage

**Affiliations:** Department of Nursing and Midwifery, Faculty of Allied Health Sciences, University of Sri Jayewardenepura, Sri Lanka; Faculty of Medicine, University of Sri Jayewardenepura, Sri Lanka

**Keywords:** Online learning, perception, preference, COVID-19, undergraduates, Sri Lanka

## Abstract

**Background:** The COVID-19 pandemic forced a rapid shift to online education worldwide, posing new challenges for students and educators in developing countries. Understanding university students’ perceptions and preferences toward online education is vital for sustaining effective learning in such contexts.

**Objective:** To assess perceptions and preferences toward online education among final-year undergraduates in selected state universities in the Western Province of Sri Lanka during the COVID-19 pandemic and to identify related benefits and barriers.

**Methods:** A descriptive cross-sectional study was conducted among 535 final-year undergraduates from five state universities. Data were collected via a structured, self-administered online questionnaire and analyzed using SPSS version 26 with descriptive statistics and Chi-square tests. A p-value of <0.05 was considered statistically significant.

**Results:** Participants’ mean age was 24 years (SD=1.3), and most were female (n=373;69.7%). The majority (n=488; 91.2%) attended online classes for the first time during the pandemic, and 83.9% used smartphones for learning. Over half (n=275;51.4%) demonstrated a good perception of online learning. Perception was significantly associated with university (p< 0.001), age (p= 0.001), and gender (p=0.002). Both live online classes and reading materials were preferred by 63.4% of respondents, while 57.8% desired 2–4 hours of daily sessions. Mobile data packs were the main internet source (n=325;60.7%). Major benefits included a comfortable environment (81.6%) and flexible scheduling (74.9%), whereas key barriers were data speed (78.5%), data limits (73.2%), and connectivity issues (68.4%). Student readiness emerged as the strongest determinant of online learning success.

**Conclusion:** Although perceptions of online education were largely positive, inadequate internet access and technological limitations constrained learning effectiveness. Strengthening digital infrastructure and equitable access are essential for sustainable e-learning in Sri Lanka.

## INTRODUCTION

The COVID-19 pandemic has drastically transformed the global education landscape, compelling higher education institutions to adopt online learning as an immediate alternative to conventional face-to-face instruction. The closure of educational institutions affected nearly 1.725 billion learners across 153 countries, representing 98.6% of the world’s student population [1]. In response, governments and universities worldwide implemented distance and online education to ensure the continuity of learning while adhering to health and safety measures. This abrupt transition marked a paradigm shift in pedagogical delivery, accelerating the integration of information and communication technology (ICT) into mainstream education [2].

Online learning, or e-learning, refers to the use of digital technology and the internet to facilitate instruction and knowledge exchange between learners and educators beyond physical classrooms [3]. It enables synchronous and asynchronous learning, promoting flexibility, accessibility, and self-paced study opportunities [4]. Globally, platforms such as Zoom, Microsoft Teams, Google Classroom, and Moodle have become integral to higher education during the pandemic [5]. However, the success of online education largely depends on multiple factors including students’ perception, motivation, readiness, and access to adequate technological infrastructure [6].

In developing countries like Sri Lanka, the rapid digital transformation posed numerous challenges, especially concerning access to reliable internet connectivity, device ownership, and digital literacy [7]. The Sri Lankan government temporarily closed all educational institutions in March 2020 to curb the spread of COVID-19 [8]. Consequently, the University Grants Commission (UGC), in collaboration with the Telecommunications Regulatory Commission of Sri Lanka (TRCSL), facilitated free data access to university learning management systems (LMS) through the Lanka Education and Research Network (LEARN) [9]. This initiative enabled students and lecturers to continue academic activities online using platforms such as Zoom, Google Meet, and university LMS portals. Nonetheless, disparities in socioeconomic background, geographic location, and institutional capacity have resulted in unequal learning experiences among students [10].

Students’ perception and preference play a crucial role in determining the effectiveness of online learning. Perception encompasses learners’ attitudes, beliefs, and satisfaction toward the online learning process, whereas preference reflects the mode, tools, and approaches they favor for optimal learning outcomes [11]. Positive perception and preference are often linked to enhanced engagement, better academic performance, and sustained motivation [12]. Conversely, inadequate technical support, poor internet connectivity, and lack of self-discipline can negatively influence students’ acceptance of online learning [13]. Studies conducted in other South Asian countries have identified similar constraints, emphasizing the need to evaluate learners’ perspectives to ensure the sustainability of online education [14,15].

In the Sri Lankan context, limited empirical research has been conducted on students’ perceptions and preferences regarding online education, particularly within the university sector. Prior to the pandemic, online teaching was minimal in state universities, and most academic programs were delivered in traditional classroom settings [16]. The sudden transition to virtual platforms during COVID-19 represented an unprecedented challenge for students and educators alike. Understanding students’ experiences, perceptions, and preferences is therefore essential for identifying the benefits, limitations, and potential improvements required in online education systems.

This study focuses on final-year undergraduates in selected state universities in the Western Province, Sri Lanka, as this group represents the most academically mature segment of the university population. Final-year students are expected to demonstrate a higher level of academic engagement and independence; thus, their experiences can offer valuable insights for institutional planning and policy formulation. The findings of this study will help identify the key determinants influencing successful online learning and contribute to developing effective digital learning frameworks for higher education in Sri Lanka.

In summary, the COVID-19 pandemic has accelerated the adoption of online learning, creating both opportunities and challenges for students and educators. While online platforms have ensured educational continuity, issues related to access, equity, and pedagogical adaptation persist. Evaluating the perceptions and preferences of university students is vital to improving the quality, inclusiveness, and sustainability of online education in Sri Lanka’s higher education system.

## METHODS

### Study Design

A descriptive cross-sectional quantitative research design was employed to assess the perception and preference for online education among final-year undergraduates in selected state universities in the Western Province, Sri Lanka, during the COVID-19 pandemic.

### Study Setting

The study was conducted among final-year undergraduates in five state universities located in the Western Province: the University of Sri Jayewardenepura, University of Colombo, University of Kelaniya, University of Moratuwa, and the University of Visual and Performing Arts. These universities were selected to represent a diverse range of academic fields including science, medicine, management, engineering, and creative arts. The Western Province was chosen because it houses the largest concentration of universities and has comparatively better internet connectivity, making it suitable for online data collection (17).

### Study Population

The study population consisted of final-year undergraduate students enrolled in the selected universities during the 2021 academic year. This group was targeted because they were the most academically advanced and had significant experience with both traditional and online learning environments. Moreover, they were heavily affected by the shift to online learning during the COVID-19 pandemic, especially in courses involving laboratory or clinical components (18).

### Inclusion and Exclusion Criteria

Inclusion criteria included final-year undergraduates from the selected universities who had participated in online learning during the pandemic and voluntarily consented to participate.

Exclusion criteria included undergraduates from private universities or other provinces, students in lower academic years, and incomplete responses.

### Sampling Technique and Sample Size

A convenience sampling method was utilized due to restrictions on physical access to campuses during the pandemic (19). The sample size was calculated using the Lwanga and Lemeshow (1991) formula for estimating proportions with a 95% confidence level and 5% margin of error, assuming a 50% expected prevalence of good perception. The minimum required sample size was 384; however, 535 completed responses were obtained to enhance accuracy and representation.

Participants were distributed as follows: University of Sri Jayewardenepura (n = 204; 38.1%), University of Colombo (n = 105; 19.8%), University of Kelaniya (n = 130; 24.3%), University of Moratuwa (n = 67; 12.5%), and University of Visual and Performing Arts (n = 28; 5.2%).

### Study Instrument

A structured, self-administered online questionnaire was developed by the researcher after a comprehensive literature review and adaptation of previously validated tools assessing e-learning perception and preference (3,5,11,14). The questionnaire, developed in Google Forms, included both closed-ended and Likert-scale questions and consisted of five main sections:

1. Sociodemographic Data – University, faculty, gender, age, and living area.
2. Online Learning Experience – Previous exposure to online classes, device used, preferred platform, and duration of participation.
3. Perception Scale – Twenty statements were rated on a five-point Likert scale (1 = strongly disagree to 5 = strongly agree). Items covered five domains: flexibility, accessibility, interaction, technical efficiency, and self-discipline. The overall perception score was computed by summing all responses, giving a maximum of 100 and a minimum of 20. The mean perception score was 76.18% (SD = 13.19). Respondents scoring above this mean were classified as having a good perception, while those below were categorized as having a poor perception (20,21).
4. Preference Scale – Focused on students’ preferred online learning modalities, including live classes, recorded materials, or blended formats, and preferred evaluation methods such as quizzes, assignments, and online exams.
5. Benefits and Barriers Scale – Designed to identify the key factors that facilitated or hindered effective online learning.

### Validity and Reliability

The questionnaire’s validity and reliability were ensured through a two-step process. Content validity was established by three academic experts in nursing and education, resulting in a Content Validity Index (CVI) of 0.92. Reliability testing using Cronbach’s alpha indicated strong internal consistency (α = 0.87) for the overall instrument, which aligns with the acceptable threshold of α ≥ 0.70 for educational research (6,22).

### Data Collection Procedure

Data were collected from July to September 2021. After obtaining ethical approval, the Google Form link was distributed to students via faculty WhatsApp groups and university mailing lists. The first section of the online form included an information sheet detailing the study objectives, ethical assurances, and contact information for the research team. Participants were required to indicate informed consent electronically before accessing the questionnaire.

All respondents received the same version of the survey with identical instructions. To maintain anonymity, personal identifiers such as names or student numbers were not collected. Data collection remained open for three weeks, and reminders were sent twice during the period to maximize response rates. This approach facilitated broad participation while adhering to COVID- 19 safety guidelines (23).

### Data Analysis

Data analysis was conducted using SPSS version 26.0. Descriptive statistics (frequencies, percentages, means, and standard deviations) were used to summarize demographic variables, perception, preferences, benefits, and barriers.

Inferential statistics, particularly the Chi-square (χ²) test, were applied to determine associations between demographic factors (university, gender, age, living area) and perception levels. A p-value of <0.05 was considered statistically significant, and results were presented with 95% confidence intervals (24). Tables and graphs were used to illustrate findings clearly.

### Ethical Considerations

Ethical approval for the study was obtained from the Ethics Review Committee of the Faculty of Medical Sciences, University of Sri Jayewardenepura (Ref: Nur/12/20). Participation was voluntary, and electronic informed consent was obtained before data collection. Confidentiality and anonymity were maintained throughout the process, and participants were informed of their right to withdraw at any time without penalty.

The study adhered to the ethical principles outlined in the Declaration of Helsinki, ensuring respect for autonomy, beneficence, and justice (25).

## RESULTS

### Demographic Characteristics of Participants

A total of 535 final-year undergraduate students participated in the study, representing five state universities in the Western Province of Sri Lanka. The mean age of participants was 24 years (SD = 1.3). The majority were female (n = 373; 69.7%), and most resided in urban or peri-urban areas (n = 428; 80.0%).

**Table 1.** Socio-demographic characteristics of participants (n = 535)

| Variable | Category | Frequency (n) | Percentage (%) |
| --- | --- | --- | --- |
| Age (years) | Mean ± SD | 24 ± 1.3 | — |
| Gender | Male | 162 | 30.3 |
|  | Female | 373 | 69.7 |
| University | University of Sri Jayewardenepura | 204 | 38.1 |
|  | University of Colombo | 105 | 19.8 |
|  | University of Kelaniya | 130 | 24.3 |
|  | University of Moratuwa | 67 | 12.5 |
|  | University of Visual and Performing Arts | 28 | 5.2 |
| Residence | Urban | 286 | 53.5 |
|  | Peri-urban | 142 | 26.5 |
|  | Rural | 107 | 20.0 |
| Primary Device Used | Smartphone | 449 | 83.9 |
|  | Laptop/Computer | 68 | 12.7 |
|  | Tablet/Other | 18 | 3.4 |
| **Internet Source** | Mobile Data Pack | 325 | 60.7 |
|  | Wi-Fi Connection | 210 | 39.3 |

### Descriptive Statistics of Key Variables Perception toward Online Learning

More than half of the participants (n = 275; 51.4%) demonstrated a good perception toward online education, while 48.6% (n = 260) had a poor perception. The mean perception score was 76.18% (SD = 13.19).

Students agreed that online learning was flexible (mean = 4.1 ± 0.9) and time-saving (mean = 4.0 ± 1.0). However, low interactivity and poor internet connectivity were commonly identified weaknesses (mean = 2.8 ± 1.1).

**Figure 1.**
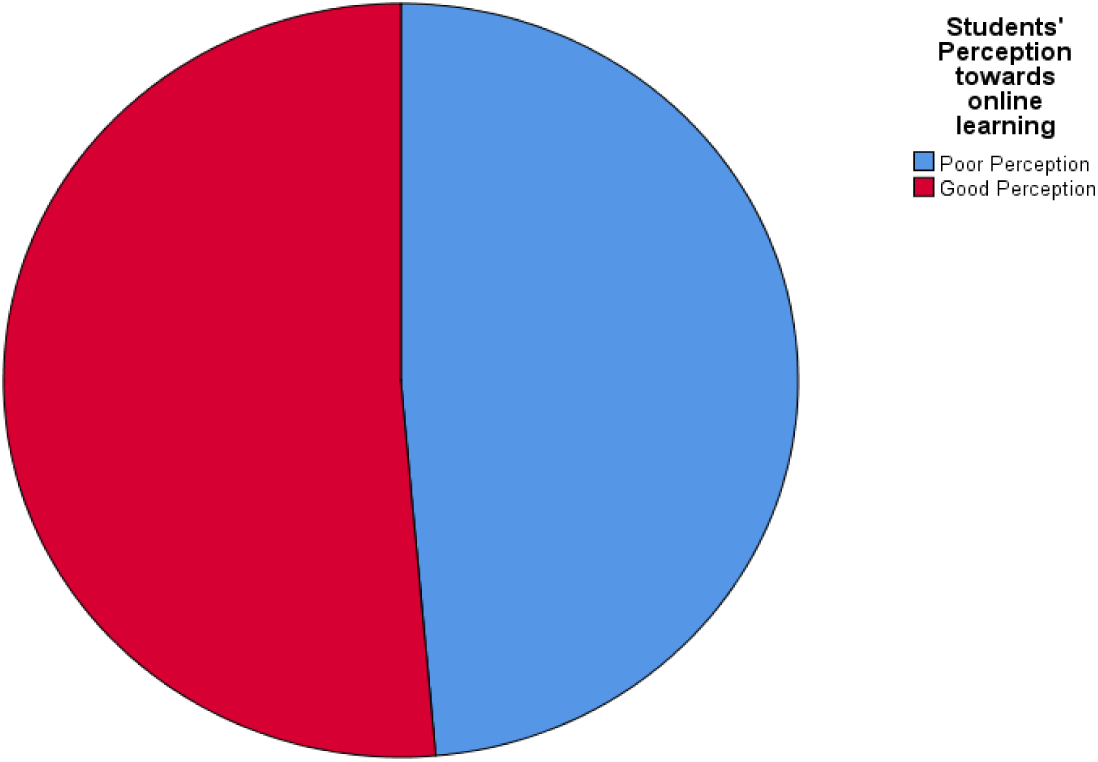
Perception Toward Online Learning among Final-Year Undergraduates (n = 535)

### Preferences for Online Learning

The majority of students (n = 339; 63.4%) preferred a combination of live online lectures and reading materials, while 23.2% (n = 124) preferred only live sessions. Most participants (n = 309; 57.8%) indicated that the ideal class duration should be between 2–4 hours per day, and 68.2% (n = 365) preferred that online classes be scheduled in the morning hours.

### Perceived Benefits and Barriers

The most frequently reported benefits of online learning were:

- Comfortable learning environment (81.6%)
- Flexible schedules (74.9%)
- Reduced travel cost (69.2%) The major barriers were:
- Low data speed (78.5%)
- Limited data capacity (73.2%)
- Poor network connectivity (68.4%)
- Lack of interaction with lecturers (65.8%)

**Table 2.** Summary of perception, preferences, benefits, and barriers (n = 535)

| Variable | Category / Indicator | Frequency (n) | Percentage (%) | Mean ± SD |
| --- | --- | --- | --- | --- |
| **Overall Perception** | Good | 275 | 51.4 | 76.18 ± 13.19 |
|  | Poor | 260 | 48.6 | — |
| **Preferred Learning Mode** | Live + Reading materials | 339 | 63.4 | — |
|  | Only Live lectures | 124 | 23.2 | — |
|  | Recorded lessons only | 72 | 13.4 | — |
| **Preferred Duration** | 2–4 hours/day | 309 | 57.8 | — |
|  | < 2 hours/day | 116 | 21.7 | — |
|  | > 4 hours/day | 110 | 20.5 | — |
| **Top Benefits** | Comfortable environment | 437 | 81.6 | $4.2 \pm 0.7$ |
| | Flexible schedule | 401 | 74.9 | $4.0 \pm 0.8$ |
| | Reduced travel cost | 370 | 69.2 | $3.9 \pm 0.9$ |
| **Major Barriers** | Low data speed | 420 | 78.5 | $2.8 \pm 1.1$ |
| | Data limitations | 392 | 73.2 | $2.9 \pm 1.0$ |
| | Poor connectivity | 366 | 68.4 | $3.0 \pm 1.0$ |
| | Lack of interaction | 352 | 65.8 | $2.7 \pm 1.2$ |

### Associations between Demographic Factors and Perception

Associations between perception level (good vs. poor) and selected demographic variables were examined using the Chi-square (χ²) test.

Significant associations were found between:

- Gender and perception (χ² = 9.73, *p* = 0.002)
- Age and perception (χ² = 7.21, *p* = 0.001)
- University attended and perception (χ² = 16.34, *p* < 0.001)

No statistically significant associations were observed with place of residence (*p* = 0.08) or primary device used (*p* = 0.22).

**Table 3.** Association between demographic factors and perception toward online learning (n = 535)

| Variable | Category | Good Perception<br>n (%) | Poor Perception<br>n (%) | $\chi^2$ | p-value |
| --- | --- | --- | --- | --- | --- |
| Gender | Male | 67 (41.4) | 95 (58.6) | 9.73 | 0.002* |
|  | Female | 208 (55.8) | 165 (44.2) |  |  |
| Age (years) | $\leq 24$ | 198 (54.8) | 163 (45.2) | 7.21 | 0.001* |
| | $> 24$ | 77 (43.3) | 97 (56.7) | | |
| University | USJ | 121 (59.3) | 83 (40.7) | 16.34 | <0.001* |
|  | UoC | 45 (42.9) | 60 (57.1) |  |  |
|  | UoK | 70 (53.8) | 60 (46.2) |  |  |
|  | UoM | 26 (38.8) | 41 (61.2) |  |  |
|  | UVPA | 13 (46.4) | 15 (53.6) |  |  |
| **Residence** | Urban | 153 (53.5) | 133 (46.5) | 3.04 | 0.08 |
|  | Rural/Peri-urban | 122 (49.4) | 127 (50.6) |  |  |
| **Primary Device Used** | Smartphone | 238 (53.0) | 211 (47.0) | 1.53 | 0.22 |
|  | Laptop/Other | 37 (45.7) | 44 (54.3) |  |  |

*Significant at $p < 0.05$ .*

## DISCUSSION

This study assessed the perception and preference for online education among final-year undergraduates in selected state universities in the Western Province of Sri Lanka during the COVID-19 pandemic. The findings revealed that slightly more than half (51.4%) of students demonstrated a good perception toward online learning, indicating moderate acceptance of digital education within the Sri Lankan higher education context. This result is comparable to studies conducted in India and Bangladesh, where 52–56% of students expressed satisfaction with online classes introduced during the pandemic (27,28).

The mean perception score of 76.18 ± 13.19 indicates a generally positive but variable outlook among students, suggesting that individual and institutional factors influence adaptability to online learning. Despite the overall favorable perception, 48.6% of students reported poor perceptions, highlighting the uneven quality and accessibility of online education across universities. Similar disparities have been documented in South Asian countries, where digital inequality and inconsistent institutional support continue to hinder effective e-learning (29).

The current study found that gender (p = 0.002), age (p = 0.001), and university (p < 0.001) were significantly associated with perception level. Female students demonstrated higher levels of positive perception (55.8%) compared with males (41.4%). This gender difference is consistent with previous findings by Ong and Lai (39), who noted that female learners often display stronger self-regulation, time management, and motivation in structured online environments. Younger students (≤24 years) also exhibited more favorable attitudes (54.8%), possibly reflecting greater technological fluency and openness to digital tools (40).

Variation among universities indicates institutional disparities in e-learning preparedness. Universities with better digital infrastructure and staff training likely provided smoother transitions to online teaching, contributing to more positive student experiences (41). Conversely, institutions with limited technological capacity may have faced difficulties in delivering consistent, interactive sessions.

The preference data showed that 63.4% of participants favored a combination of live classes and reading materials, while 23.2% preferred live-only sessions. This preference for blended learning aligns with global literature suggesting that combining synchronous and asynchronous formats enhances flexibility and engagement (30,31). Most students (57.8%) recommended a daily duration of 2–4 hours for online learning, consistent with studies highlighting that shorter, interactive sessions are more effective in maintaining attention and preventing screen fatigue (31).

In terms of benefits, the majority of participants cited a comfortable learning environment (81.6%), flexible scheduling (74.9%), and reduced travel costs (69.2%) as major advantages. These findings mirror those reported in Malaysia, the Philippines, and India, where online learning was perceived as convenient, cost-effective, and accessible (32–34). The comfort of home learning may also have reduced academic stress and allowed better time management. However, self-discipline and digital literacy remain crucial factors influencing academic success in remote education (35).

The major barriers identified were low data speed (78.5%), limited data capacity (73.2%), and poor connectivity (68.4%), followed by lack of interaction with lecturers (65.8%). These issues are consistent with findings from other developing countries, where unreliable internet access and inadequate bandwidth significantly affect student participation (29,36). In Sri Lanka, although initiatives like the Lanka Education and Research Network (LEARN) provided free access to university Learning Management Systems, limitations in network coverage and affordability persisted (37).

Reduced interaction and feedback emerged as key concerns, emphasizing the importance of social and teaching presence in online education. According to Garrison’s Community of Inquiry Framework, meaningful learning outcomes depend on continuous instructor engagement and peer collaboration (38). Lack of interaction may lead to reduced motivation and superficial learning, underscoring the need for pedagogical innovations that encourage communication through discussion forums, group projects, and virtual simulations.

Overall, these findings align with global trends showing that while online learning offers flexibility and continuity, it also exposes inequalities in access and digital readiness (33,34,36). Universities that adopt blended learning approaches tend to report higher student satisfaction and improved performance outcomes (43). For Sri Lankan universities, the results underscore the urgent need to strengthen digital infrastructure, provide affordable internet access, and enhance lecturers’ competence in online teaching methodologies.

Despite its strengths, this study has limitations, including the use of convenience sampling and self-reported data, which may introduce bias. However, the inclusion of students from five major universities enhances external validity and provides a realistic picture of the higher education landscape in Sri Lanka.

In conclusion, the study found that Sri Lankan undergraduates hold generally positive perceptions toward online learning, recognizing its convenience and flexibility. However, persistent barriers related to connectivity, interaction, and institutional preparedness continue to constrain its effectiveness. Improving technological infrastructure, promoting faculty training, and adopting sustainable blended learning models will be crucial for ensuring equitable and high-quality online education in the post-pandemic era.

## LIMITATIONS

This study has several limitations. As a cross-sectional survey conducted using a convenience sampling technique, it may not fully represent all final-year undergraduates across Sri Lanka. Data were self-reported, which may introduce recall or response bias. The study focused only on selected universities in the Western Province, limiting generalizability to other regions.

Additionally, the quantitative design restricted exploration of deeper qualitative insights into student experiences. Despite these limitations, the inclusion of five universities and a relatively large sample size (n = 535) provides a robust overview of online learning perceptions during the COVID-19 pandemic.

## CONCLUSION

This study revealed that over half (51.4%) of final-year undergraduates in selected state universities in Sri Lanka had a good perception toward online learning during the COVID-19 pandemic, with a mean perception score of 76.18 ± 13.19. Students valued flexibility (74.9%), comfort (81.6%), and cost-effectiveness (69.2%), yet faced major barriers such as low data speed (78.5%) and limited connectivity (68.4%). Significant associations between perception and gender (p = 0.002), age (p = 0.001), and university (p < 0.001) highlight disparities in access and institutional readiness.

The findings underscore the need for enhanced digital infrastructure, affordable internet access, and lecturer training to improve online teaching quality and student engagement. Adopting a blended learning approach, integrating both synchronous and asynchronous elements, could provide a balanced and sustainable educational model for the post-pandemic era. Strengthening policy support, technological equity, and pedagogical innovation will be essential for ensuring inclusive, high-quality online education in Sri Lanka’s higher education system.

## Data Availability

Data supporting the findings are available from the corresponding author upon reasonable request.

## Acknowledgments

The author sincerely thanks all participating students and university staff for their cooperation during data collection.

## DECLARATIONS

### Author Approval

All authors have reviewed and approved the final manuscript.

### Data Availability Statement

Data supporting the findings are available from the corresponding author upon reasonable request.

### Ethical Approval

Approved by the Ethics Review Committee, Faculty of Medical Sciences, University of Sri Jayewardenepura (Ref: Nur/12/20).

